# The Impact of Self-Management-Based Care Interventions on Quality of Life in Type 2 Diabetes Mellitus Patients: A Philosophical Perspective

**DOI:** 10.1101/2022.06.27.22276988

**Authors:** Fadli

## Abstract

Type 2 diabetes mellitus is caused by the disruption of insulin secretion and resistance. One aspect that plays an important role in this disease is self-management education. Good self-care behavior facilitates controlled diabetes management and prevents complications as well as ensures a better life quality. This literature aims to study the philosophy of diabetes self-management based care interventions to improve the quality of life viewed from philosophical perspectives. A literature search was performed on Scopus, PubMed, ProQuest, and Science Direct using keywords including type 2 diabetes mellitus, diabetes self-management and quality of life. The inclusion criteria are peer-reviewed articles in English that discuss diabetes self-management and quality of life. Articles published within the last five years (2017-2021). Research such as a randomized controlled trial (RCT) and full text method. Based on the philosophy study self care interventions, eight articles showed that self-management interventions provide significant effectiveness for lifestyle changes and self-care for type 2 diabetes mellitus patients. Eight articles were selected based on an axiological philosophical study approach, six of which discussed self-management interventions’ effect on self-care behavior, and two measured the quality of life of type 2 diabetes mellitus patients. Almost all the articles stated an increase in self-care behaviors and quality of life after receiving self-management interventions. Successful diabetes self-management depends on individual self-care activities to control symptoms presented. Furthermore, regular self-management activities prevent complications from arising. Therefore, patients’ compliance with diabetes self-management is needed to improve their life quality.

## 1. INTRODUCTION

Diabetes Mellitus (DM) is a chronic non-communicable (Mosleh *et al*., 2017) metabolic disease with a disturbance in the hormone insulin, which functions in maintaining the body’ s homeostasis by decreasing blood sugar levels (American Diabetes Association, 2017). Type 2 diabetes mellitus is one of the most common types experienced by the population (Joyce & Jane, 2014). This is a chronic disease and global health problem, affecting approximately 422 million people worldwide (Moura et al., 2019).

Based on data from the International Diabetes Federation (IDF) in 2021, it is estimated that 537 million people, i.e. approximately 1 in 10 people suffer from diabetes worldwide.

The data is estimated to reach 643 million people in 2030 and the total cases are predicted to increase by 783 million people in the age range of 20-79 years in 2045 provided no intervention is carried out (IDF, 2021). Globally, Indonesia is currently ranked as the fifth country with the highest diabetes mellitus cases, namely 19.5 million, and is estimated to increase by 28.6 million people in 2045 (Perkeni, 2021).

In addition to the increasing incidence of cases, DM also causes many acute and chronic complications. The acute complications include diabetic ketoacidosis, nonketotic hyperosmolar, and hypoglycemia, while macroangiopathy, microangiopathy, and neuropathy are chronic (Papachristoforou, Lambadiari, Maratou, & Makrilakis, 2020). Once any of these occur, costs of survival are increased, and life quality becomes affected. A study in Palestine showed that almost 34% of DM patients experienced a poor life quality (Tietjen et al., 2021). Meanwhile, several sources reported that DM patients in Indonesia averagely had a decrease in life quality. According to Umam, Solehati, and Purnama (2020), patients’ life quality was mostly 63.7% in the moderate category. Based on the physical, psychological, social relation, and environmental domains, 61.5%, 60.4%, 58.2%, and 53.8% were in the moderate category respectively.

Once self-management is not controlled, patients experience DM all through their lifetime, thereby leading to a great effect on the life quality. Good and bad self-management in the patients is influenced by several factors, namely age, gender, education level, length of suffering, knowledge, self-efficacy, diabetes stress, and family support (Lin et al., 2017; Ningrum, Alfatih, & Siliapantur, 2019). Effective self-management in patients is important to improve goal achievement during management. Non-adherence to treatment hinders the regulation of blood sugar levels, leading to poor glucose control (Hsu, Lee, & Wang, 2018).

According to Kurniawan, Sari, and Aisyah (2020), out of 123 respondents, 62.6% had low self-management on blood sugar monitoring indicators. Meanwhile, a Chinese study showed a moderate category of self-management behavior in 50.4% of diabetes patients, and 33.6% had low self-management (Qi et al., 2021). Considering this, some patients do not still know about self-management in-depth and correctly. Various interventions to improve patients’ self-management are carried out in the form of diabetes mellitus self-care and self-management education, but no optimal results are obtained yet, and many people have not shown independence in managing their disease. To manage the disease effectively, a spiritual approach is needed to control the patients’ emotions and self-concept. Furthermore, the increase of families’ knowledge and skills in helping patients overcome their disease problems is necessary to achieve life quality improvement. Therefore, This literature aims to study the philosophy of diabetes self-management based care interventions to improve the quality of life viewed from philosophical perspectives.

## 2. METHOD

The study method used an integrative literature review. This review proposes an self-management-based care interventions on quality of life in type 2 diabetes mellitus patients. We systematically searched Scopus, PubMed, ProQuest, and Science Direct. Search using various combinations of keywords with the help of Boolean operators, including: “type 2 diabetes mellitus” AND “diabetes self-management “ AND “quality of life”, combined as MESH term and keyword. The inclusion criteria applied in this study are peer-reviewed articles in English that discuss diabetes self-management and quality of life. Articles published within the last five years (2017-2021). Research studies are carried out in various areas that specifically examine self-management-based care interventions on quality of life in type 2 diabetes mellitus patients. This study is a quantitative study with a randomized controlled trial (RCT) and full text method. The first author performs an initial database search and articles for review. We used the PRISMA Flowchart 2009 (Moher et al., 2010) to record the article review and inclusion process (see Figure 1). An initial search of four databases yielded 1.137 results. After that, we collected all articles and removed duplicated articles. The source was excluded by title and abstract if it was not a peer-reviewed research study or related to self-management-based care interventions on quality of life in type 2 diabetes mellitus patients in nursing scope. The next step was to narrow the selection of articles based on the year of publication and the research contexts. After the remaining articles were assessed for significant quality of life in type 2 diabetes mellitus patients findings, 8 papers were selected for final inclusion **(Figure 1)**.

**Figure 1.**
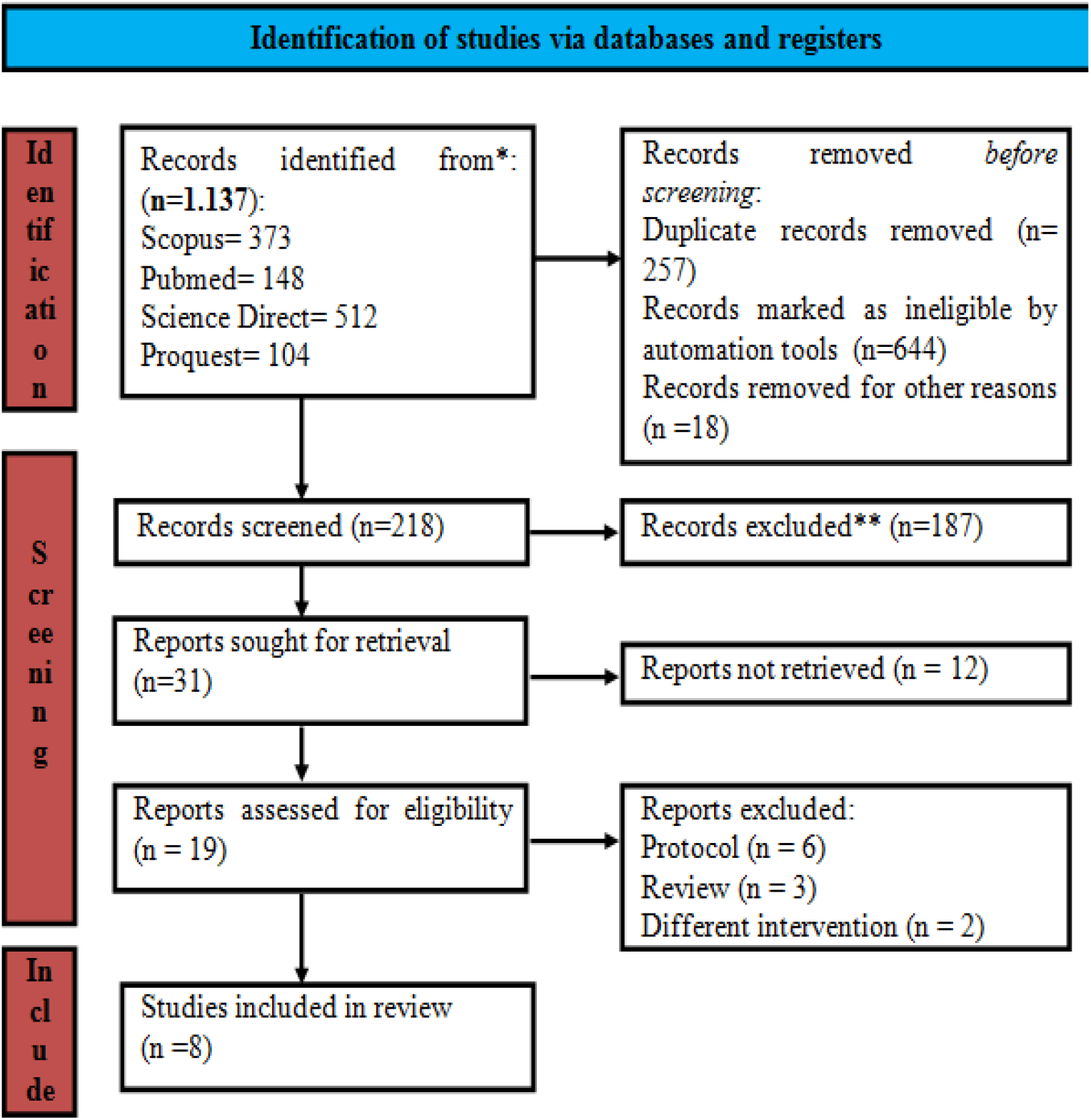
PRISMA Flowchart of Literature Search and Screening Process

## 3. RESULT AND DISCUSSION

### a. Philosophy of care intervention based on diabetes self-management from an Ontology perspective

Type 2 diabetes mellitus is a chronic non-communicable disease due to disturbances in the insulin hormone which functions in maintaining the body’ s homeostasis by decreasing blood sugar levels (American Diabetes Association, 2017), and is a global health problem affecting approximately 422 million people worldwide (Moura et al., 2019). In addition to the increasing cases, DM also causes many acute and chronic complications. The acute complications include diabetic ketoacidosis, nonketotic hyperosmolar, and hypoglycemia, while macroangiopathy, microangiopathy, and neuropathy are chronic (Papachristoforou *et al*., 2020). Once any of these occur, survival costs are increased, and life quality becomes affected.

A study in Palestine stated that almost 34% of DM patients experienced a poor life quality (Tietjen *et al*., 2021). Meanwhile, several results showed that the diabetes mellitus patients in Indonesia averagely had a decrease in life quality. Based on the ontology approach study, the influencing factors are education level, knowledge, family support, income, medication adherence, and disease complications (John *et al*., 2019). According to Karami *et al*. (2021), type 2 DM patients’ life quality is influenced by age, gender, education, complications, duration, control of blood sugar levels, social support, and therapy/medication. Furthermore, the life quality assessment can be carried out by examining patients’ metabolic control results using parameters such as HbA1c (Bekele *et al*., 2021), aimed at being an indicator that diabetes has been well controlled to improve their life quality. Therefore, it is necessary to establish indicators that form self-care behavior to facilitate the intervention process to improve patients’ life quality.

The application of self-management is one aspect that plays an important role in the management of type 2 diabetes mellitus, including diet regulation, physical activity/exercise, blood sugar monitoring, compliance with medication consumption, and self/foot care (Hidayah, 2019). Effective self-management in patients is important to improve the achievement of goals in DM management. Non-adherence to diabetes medication hinders the regulation of blood sugar levels, leading to poor glucose control (Hsu, Lee, and Wang, 2018). Therefore, patients’ compliance with diabetes self-management is needed to improve their life quality. Successful diabetes self-management depends on individual self-care activities to control symptoms presented. Furthermore, regular self-management activities prevent complications from arising (Pereira *et al*., 2020).

### b. Philosophy of care intervention based on diabetes self-management from an Epistemological perspective

Knowledge is one of the supporting factors in carrying out daily self-care, which its sufficiency helps a person to understand the body’ s present condition and become capable of proper self-management to achieve a constant healthy lifestyle and controlled blood glucose. According to Orem, Self-care deficit theory focuses on each individual’ s ability to perform self-care, defined as the practice of activities that individuals initiate and perform on their own behalf in maintaining life, health, and well-being. The self-care or self-care deficit theory of nursing is composed of three interrelated theories: (1) the theory of self-care, (2) the self-care deficit theory, and (3) the theory of nursing systems, which is further classified into wholly compensatory, partially compensatory and supportive-educative (**Figure 2**).

**Figure 2.**
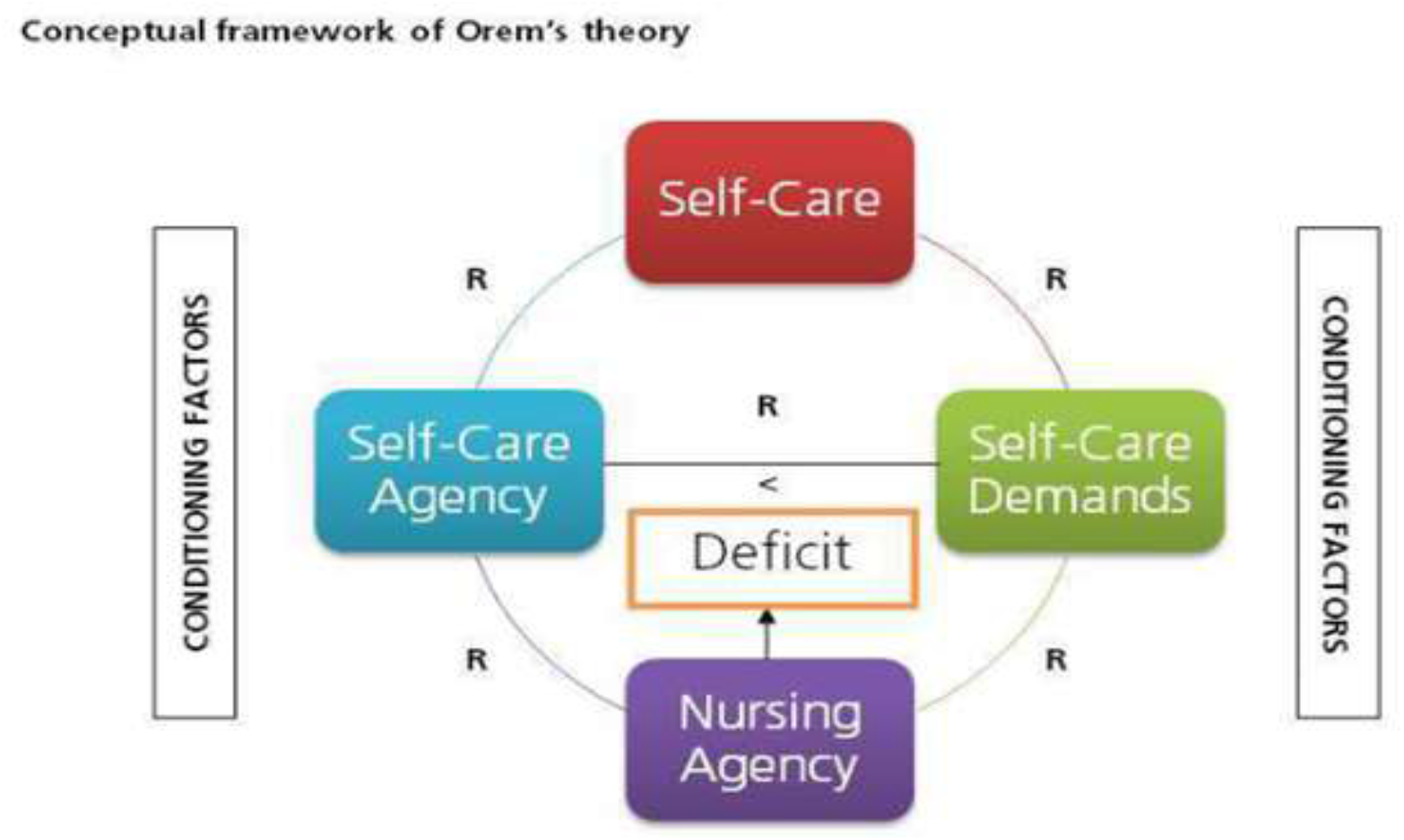
Orem’ s Self-Care Theory

Nursing Agency is a complex property or attribute of people educated and trained as nurses that enables them to act, know, and help others meet their therapeutic self-care demands by exercising or developing their own self-care agency.

The diabetes self-management education implementation adapted from the management strategy of the self-management support theory developed by Glasgow *et al*. (2003) is called the Five A’ s Model of Self-Management Support, namely, assessment, advise, agree, assist, and arrange **(Figure 3)**. Diabetes self-management education based Self-Management Support provided by nurses is expected to enhance patients’ self-care and their behavior, thereby leading to an increase in life quality and glycemic control. According to Bekele *et al*. (2021), diabetes self-management education is effective in reducing HbA1c in type 2 diabetes mellitus patients and there are differences in the results obtained by Cunningham *et al*., (2018) which showed diabetes self-management education and HbA1c had an insignificant effect, while life quality had a significant effect. Therefore, diabetes self-management education optimizes metabolic control, prevents complications, and improves the life quality of type 2 diabetes mellitus patients.

**Figure 3.**
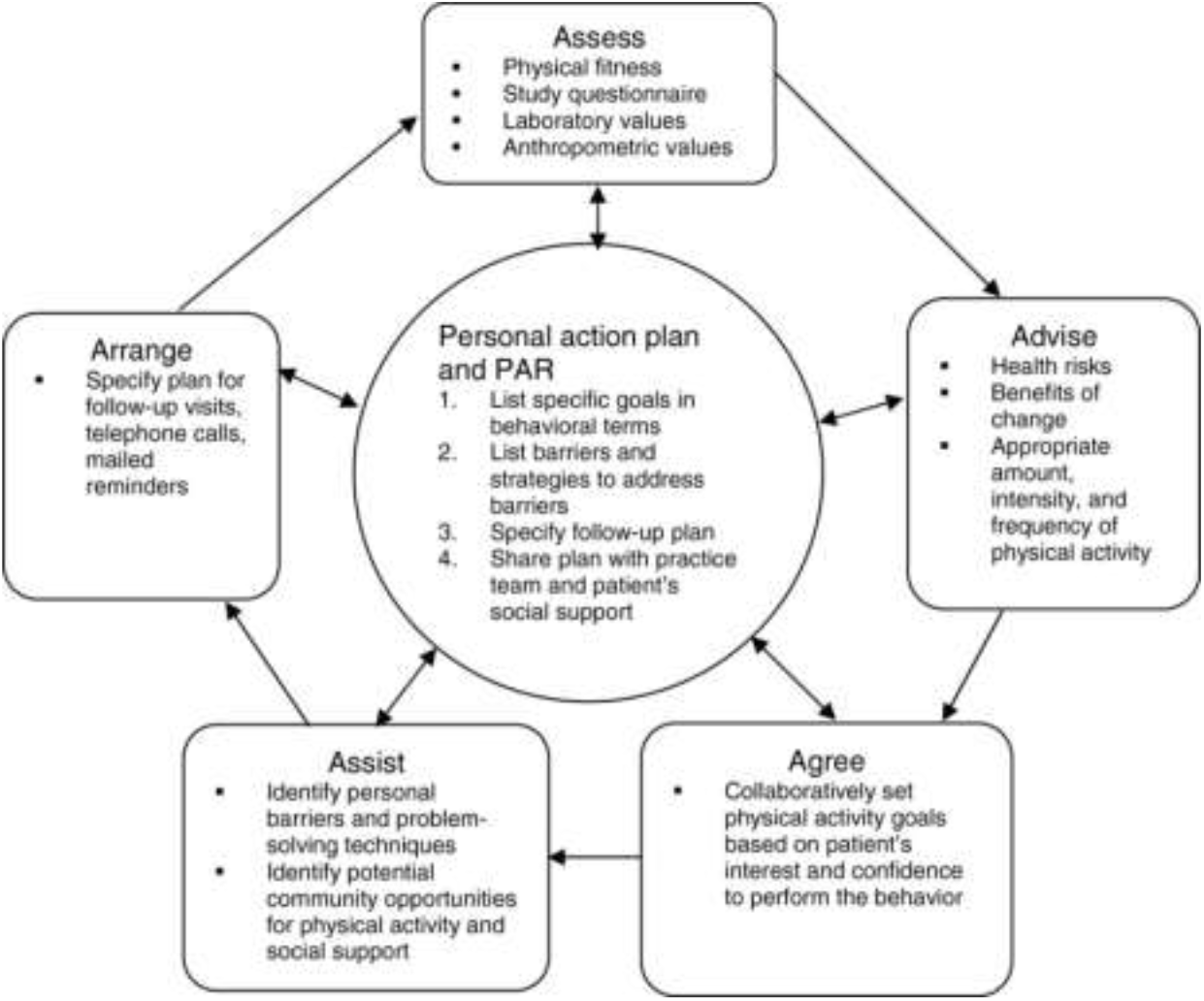
Self-Management Theory (Glasgow et al., 2003)

### c. Philosophy of care intervention based on diabetes self-management from an Axiological perspective

Based on the literature review of several studies (Table 1), Diabetes Self-Management Education (DSME) is the effective method used in improving the health status and life quality of type 2 DM patients with an epistemological approach (Rondhianto, Kusnanto, & Melaniani, 2018), (Hailu, Moen, & Hjortdahl, 2019), (Rasoul et al., 2019), and (Okafor et al., 2021). DSME is an ongoing process carried out to facilitate the knowledge, skills, and ability of DM patients to perform self-care (Hailu, Moen, and Hjortdahl, 2019). Another study used the Patient-Centered Self-Management Empowerment Intervention (PCSMEI), group-based self-management support, and social support-based self-management behavior program (Cheng et al., 2019; Puffelen, Rijken, Heijmans, Nijpels, & Schellevis, 2019; and Qi et al., 2021). This is the modification result of the Diabetes Self-Management Education (DSME) method.

**Table 1.**
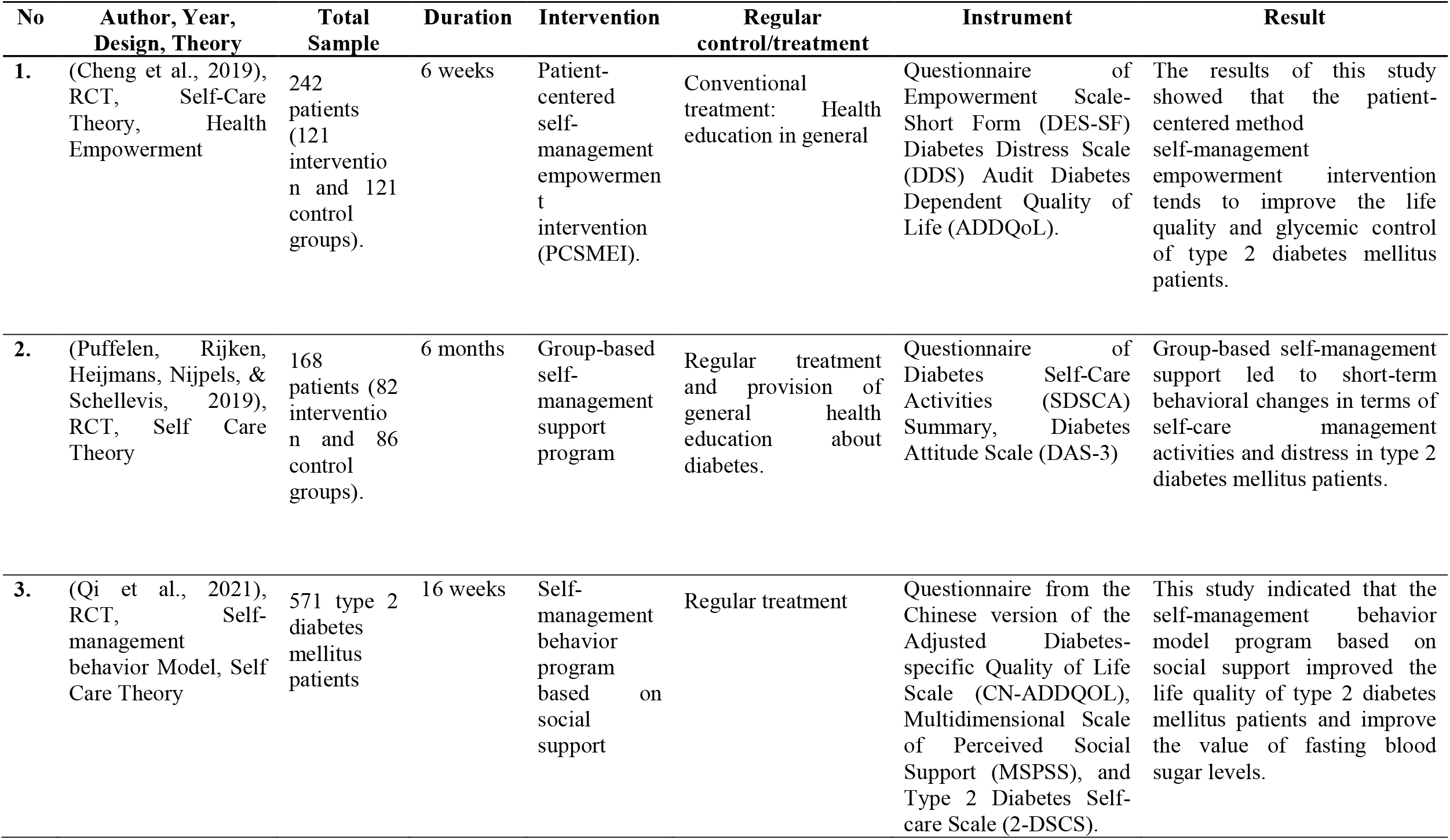
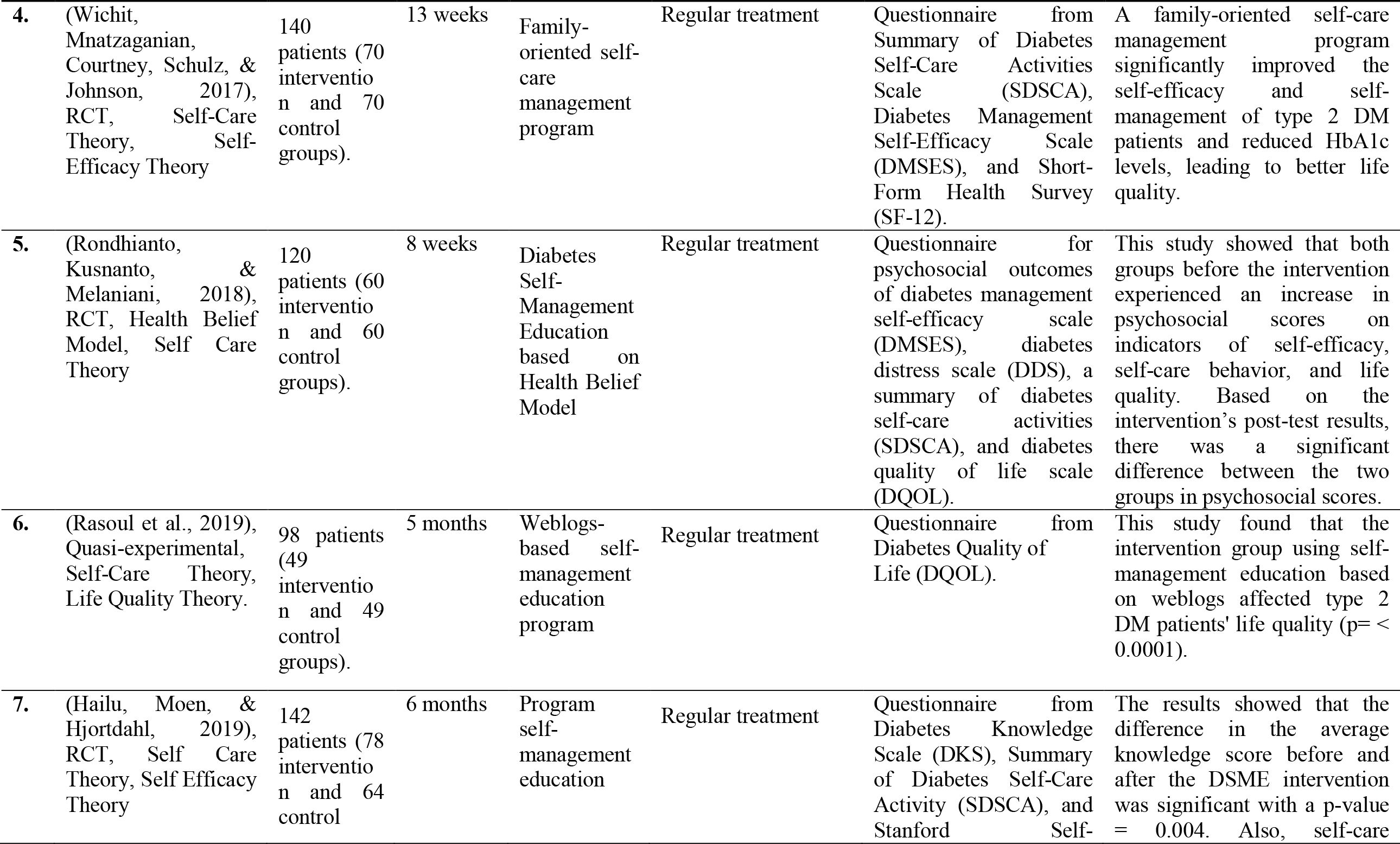
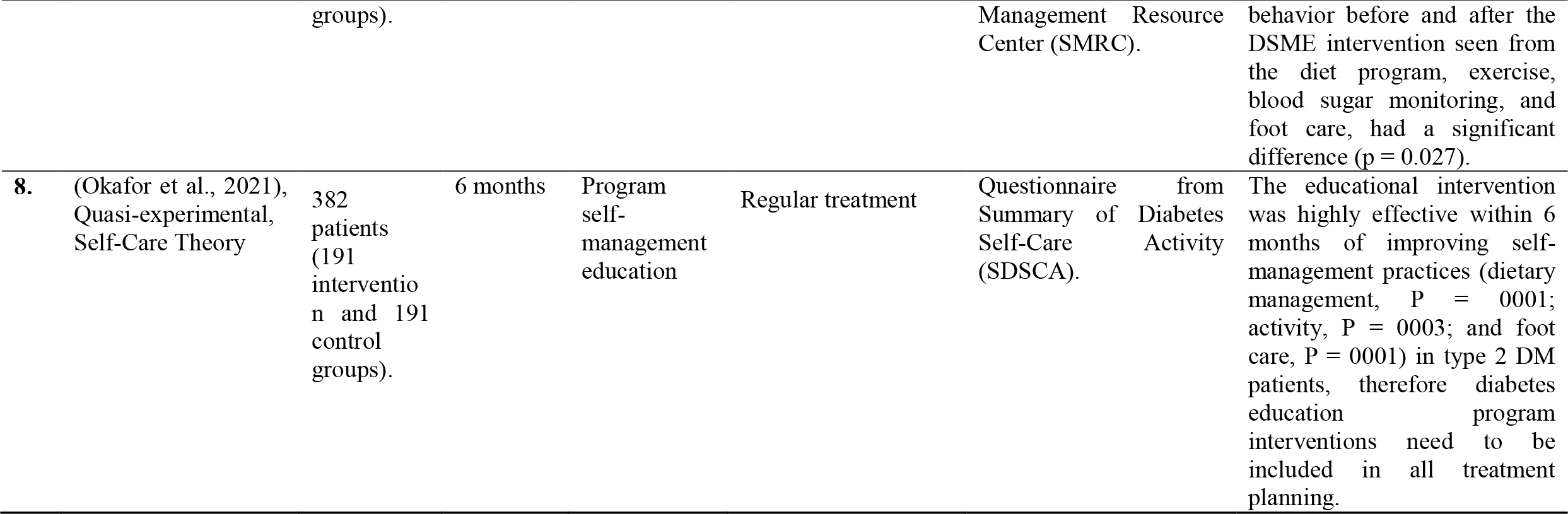
The results of the review of articles based on the Axiological perspective approach (n=8)

Based on the axiological studies, almost all studies discussed in this literature review stated that self-management tends to improve the life quality of diabetes mellitus patients. In this case, self-management ought to be an important concern for health professionals because it can be a reference for the success of an action/intervention or therapy. Furthermore, patients experience DM all through their lifetime once self-management is not controlled, leading to a great influence on the life quality (Lin *et al*., 2017). A family-oriented self-care management program significantly enhances the self-efficacy and self-management of type 2 diabetes mellitus patients, as well as reduces the HbA1c levels, thereby improving their life quality (Wichit, Mnatzaganian, Courtney, Schulz, & Johnson, 2017).

Referring to the studies reviewed, some patients do not still know about diabetes self-management in-depth and correctly. Various interventions to improve the self-management of patients are carried out in the form of diabetes mellitus self-care and self-management education, but the results are not yet optimal and many people have not shown independence in managing their disease (Hailu, Moen, and Hjortdahl, 2019). To manage the disease effectively, a spiritual approach is needed to control patients’ emotions and self-concept. Furthermore, the increase of families’ knowledge and skills in helping patients overcome their disease problems is necessary to achieve life quality improvement.

## 4. CONCLUSION

This literature review discusses the effectiveness of self-management interventions in type 2 diabetes mellitus patients with several parameters, but similarities across the literature refer to life quality. The implementation of self-management plays an important role in type 2 DM management, including diet regulation, physical activity/exercise, blood sugar monitoring, drug consumption compliance, and self/foot care. The success of diabetes self-management depends on individual self-care activities to control the symptoms presented, therefore regular self-management activities tend to prevent complications. Various interventions to improve the self-management of patients are carried out in the form of diabetes mellitus self-care and diabetes self-management education, but the results are not yet optimal and many people have not shown independence in managing their disease. It is expected that the development of self-management coaching interventions can be integrated through different theoretical approaches, namely the spiritual approach, which is needed to control patients’ emotions and self-concept. Furthermore, the increase of families’ knowledge and skills in helping patients overcome their disease problems is necessary to achieve life quality improvement.

### IMPLICATIONS OF THE RESULTS ON NURSING PRACTICE

This literature review study is expected to be used as an input for nursing science, specifically medical-surgical nursing in determining appropriate interventions for type 2 DM patients. The right intervention selection supports the process of improving patients’ health status, particularly in meeting their basic needs, thereby leading to life quality improvement.

## Data Availability

All data produced in the present work are contained in the manuscript

## ACKNOWLEDGEMENT

The authors thank to the Faculty of Nursing Universitas Airlangga for the facilities in this study.

